# Model-based optimization of controlled release formulation of levodopa for Parkinson’s disease

**DOI:** 10.1101/2023.03.17.23287393

**Authors:** Yehuda Arav, Assaf Zohar

## Abstract

Levodopa is the current standard of care for Parkinson’s disease, but chronic therapy has been associated with motor complications. Maintaining sustained and constant blood concentrations may reduce these complications, but designing a controlled release formulation (CRF) that delivers such a profile is challenging. To facilitate the development of such formulation, we developed and validated a physiologically based mathematical model. Analysis of experimental results using the model shows that levodopa is well absorbed along the entire small intestine and that levodopa in the stomach causes fluctuations during the first 3 hours after administration. Based on these insights, we used the model to develop guidelines for an optimal CRF for different stages of PD. Such formulation is expected to produce steady concentrations and prolong therapeutic duration by 50% over a common CRF with a lower dose, thereby increasing the patient’s compliance with the dosage regime.

## Introduction

Parkinson’s disease (PD) is a chronic neurodegenerative disorder caused by the progressive loss of dopamine-producing neurons in the brain (Groger, 2014). The main symptoms of PD include resting tremor, rigidity, and slow movement (bradykinesia)^1^. While there is no known cure for PD, symptom management can help improve the patient’s quality of life^2^.

The most effective drug to control the symptoms of PD is levodopa, and oral delivery is the preferred route for its administration^1,3^. However, levodopa’s commercially available oral formulations produce pulsatile, non-physiological concentrations of levodopa in the blood^4^, and relief from PD symptoms for up to 4 hours^5^. Over time, two significant problems can arise. First, the pulsatile concentrations of levodopa in the blood are considered to be one of the key factors that trigger the development of motor complications^1,6,7^ that emerge in about 50% of the patients after five years of treatment^1,3,8,9^. Second, the clinical regime may require as much as 5-10 daily doses, given at specific timing^10,11^. Adhering to such a regime is difficult, and consequently, only 27 *−* 38% of the doses are taken at the correct timing.

Developing a new oral formulation of levodopa that produces sustained, smooth blood concentrations could address the limitations of current formulations^3^. A first step toward achieving this goal is to find the optimal dose and release rate that would produce sustained therapeutic concentrations of levodopa in the blood. That is, identify the lowest possible dose that would maintain stable therapeutic concentrations of levodopa in the blood for the longest time. Physiologically based pharmacokinetic modeling is a valuable approach to optimize drug absorption^12–17^. This approach involves modeling the physiological processes following oral administration using time-dependent differential equations, either partial or ordinary. Solving these equations makes it possible to quantify the contribution of different processes to the overall absorption process and consequently find optimal solutions.

This work aims to suggest guidelines for developing an optimal oral formulation of levodopa. To accomplish this goal, we developed and validated a physiologically based mathematical model that describes the kinetics of levodopa absorption from its oral ingestion until it reaches the blood. Using the mathematical model, we were able to predict the mean concentration-time profile of levodopa in the blood based on the properties of the formulation (e.g., the dose and release rate) and to quantify the contribution of the physiological processes to the overall absorption process. The quantitative understanding of the absorption process allowed us to find the properties of a putative oral formulation expected to extend the time to deliver steady blood concentrations.

### Physiologically based mathematical model

In the following, we present a mathematical model for the absorption of levodopa following its administration in a controlled release formulation (CRF), i.e., a formulation that releases the drug over an extended period. We focus on CRF since such formulations are expected to prolong the absorption and produce smooth concentrations of levodopa in the blood^1,18^. However, clinical trials with commercially available CRFs, such as Sinemet CR, have not demonstrated a reduction in motor complications, which was attributed to erratic absorption that results in a pulsatile concentration of levodopa in the blood^3,4^. Hence, our first goal will be to use the model to understand the reason for their failure. With this understanding, we hope to suggest the properties of a putative optimal CRF. Before we use the model, it is necessary to ascertain that it can predict the blood concentrations of levodopa after oral administration of a CRF. Hence, we test it by comparing its predictions to experimental results. In particular, we compare the model predictions for the blood concentrations of levodopa following administration of different CRFs to healthy and parkinsonian individuals to the experimental blood concentrations taken from Arav^16^, Flashner et al.^19^, and Hammerstad^20^.

### Modeling the oral absorption of controlled release levodopa

Figure 1a depicts the route levodopa undergoes after oral administration in a CRF. Following ingestion, the CRF passes through the mouth cavity and the esophagus and reaches the stomach^21,22^.

**Figure 1.**
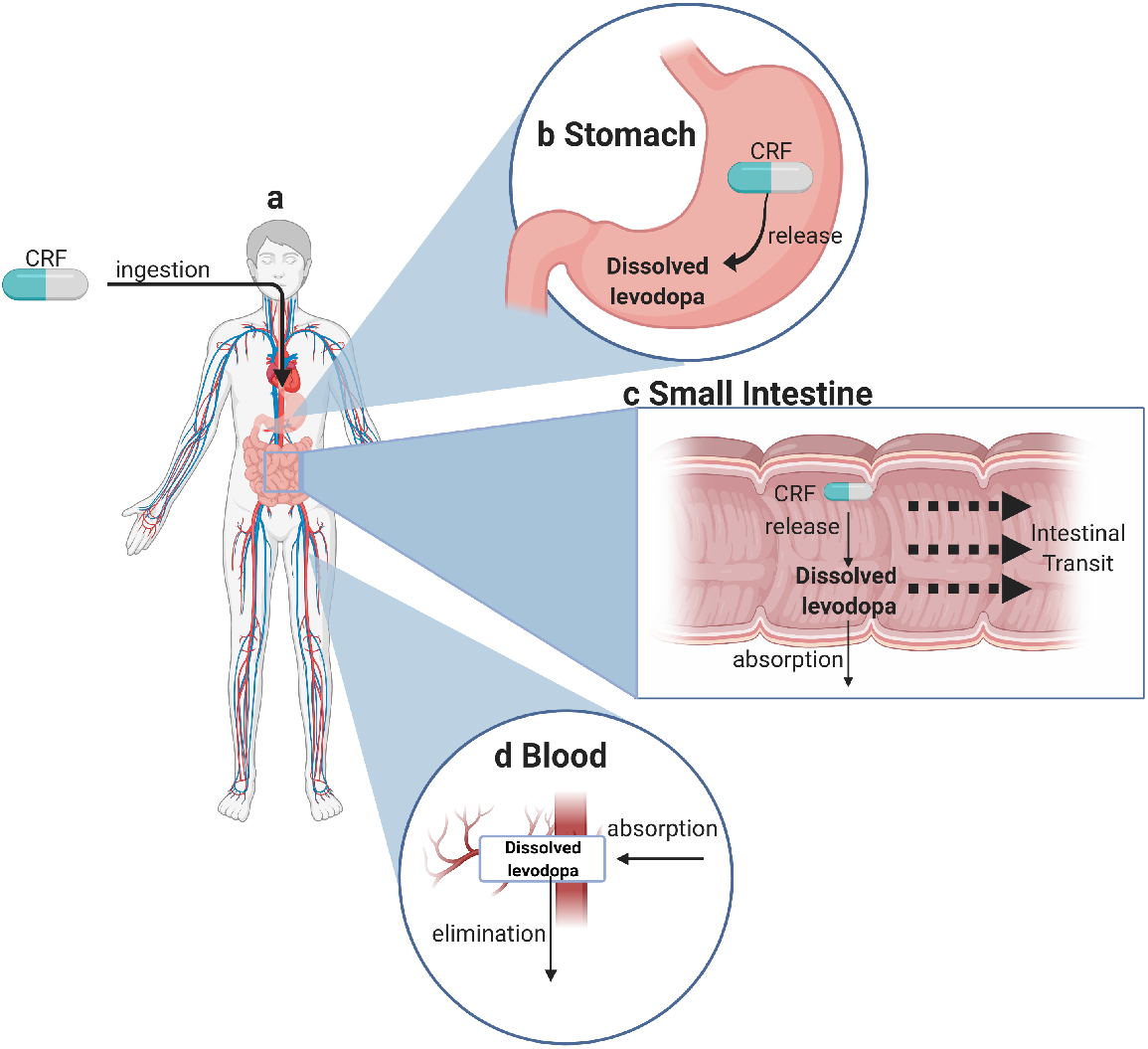
**(a)** The route of levodopa in the gastrointestinal tract following oral administration in CRF until it reaches the blood. **(b)** The processes that occur in the stomach. **(c)** The processes that occur in the small intestine. **(d)** The processes that occur in the blood. Created with BioRender.com.

#### The Stomach

A bean-shaped muscular bag that acts as a reservoir and regulates the transfer to the small intestine (SI). Figure 1b illustrates the processes that occur in the stomach. Upon reaching the stomach, the CRF begins to release levodopa. The amount of levodopa in the CRF is described by the following ordinary partial equation,

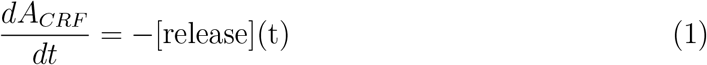

Where *A*_*CRF*_ is the amount of levodopa in the CRF. The term [release] represents the rate at which levodopa is released from the CRF, as described in Equation 9 in the Methods. The CRF remains in the stomach for a period of time before it passes into the small intestine (SI).

The levodopa that was released is rapidly dissolved^23,24^, and emptied continuously to the SI. That is, the time scale of the dissolution process is much shorter than the time scale of the other processes in the stomach (see Methods for details). Therefore, we consider the dissolution process as an instantaneous process. The concentration of dissolved levodopa in the stomach is determined by the release rate from the CRF while it is in the stomach and the transfer from the stomach to the SI (stomach emptying). After the CRF passes to the SI, the *C*_*st*_ is determined only by the stomach emptying. Due to the stomach’s shape and size, we consider the stomach as a well-mixed compartment. The ordinary partial equation gives the temporal change in the concentration of dissolved levodopa,

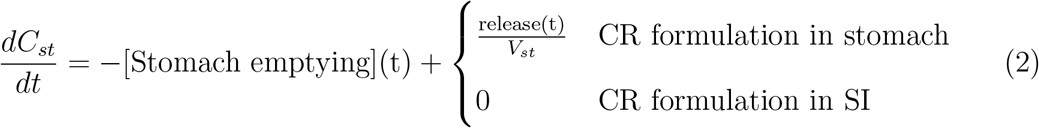

The term [Stomach emptying] represents the transfer rate of the dissolved levodopa to the SI (as described in Equation 10 in the Methods), and *V*_*st*_ is the volume of the stomach. Dissolved levodopa can cause erratic gastric emptying^25–28^. Following Robertson et al.^26^, this phenomenon was described as a period in which there is no transfer to the stomach (i.e. the [Stomach emptying] term equals 0 during that time). See the Methods section for further details.

#### The Small Intestine (SI)

A long (280cm) and narrow tube where the majority of drug and nutrient absorption occurs^13,22^. It is anatomically divided into three distinct regions: Duodenum (20cm long), Jejunum (104cm long) and the Ileum (156cm long)^13^. Three structures amplify the surface area in contact with the intestinal fluid: Large folds, finger-like projects (villi), and additional smaller protrusion (microvilli)^13,22^. The large folds are typically found between the mid-duodenum to mid-ileum, and their size and density decrease along the SI. The amplification of the Villi, finger-like projections, and the microvilli to the surface area of the SI also decreases along its length.

The processes that take place in the SI are depicted in Figure 1c. The dissolved levodopa and the CRF, which has been transferred from the stomach to the SI, are propelled toward the end of the SI through the contraction of its walls^29^. In the SI, the CRF continues to release levodopa at a similar rate to that in the stomach, which quickly dissolves^16,23^. The Large Neutral Amino Acid (LNAA) transporters actively transport the dissolved levodopa into the bloodstream^23,30,31^.

The SI is described as a continuous tube with spatially varying properties, such as the effective surface area, similar to several previous studies ^12,13,16^. The concentration of the dissolved levodopa along the SI is influenced by the stomach-emptying rate of dissolved levodopa, the release rate from the CRF, uptake to the blood by the LNAA transporters and the intestinal transit. The partial differential equation determines the temporal and spatial distribution of the dissolved levodopa in the SI,

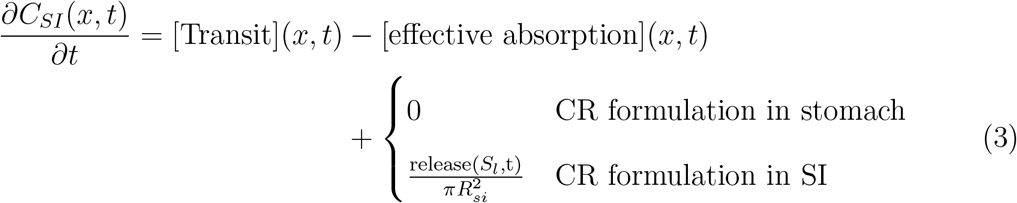

Where *x* is the location along the SI, *C*_*SI*_(*x, t*) is the concentration of dissolved levodopa along the SI, *S*_*l*_ is the location of the CR formulation along the SI, and *R*_*si*_ is the radius of the SI. The [Transit] term represents the propulsion and longitudinal mixing along the SI^12^ (as described in Equation 11 in the methods), the [effective absorption] term represents the effective uptake of levodopa by the LNAA transport system (as described below), and the [release] term represents the amount that was released from the CRF at time *t* (calculated with Equation 1) in location *S*_*l*_ (see Equation 4 below). The boundary conditions of Equation 3 at *x* = 0 accounts for the stomach emptying, and at *x* = *L*, where *L* is the length of the SI, for the emptying of the intestinal content from the ileum to the large intestine. See Methods for further details on Equation 3.

The location of the CRF along the SI is determined by the transit velocity in the SI,

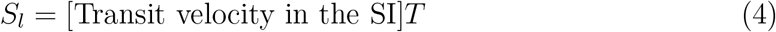

Where *T* is the elapsed time from the passage of the formulation from the stomach to the SI, the amount of levodopa in the CRF is calculated using Equation 1.

It is generally thought that levodopa is absorbed only in the upper third of the SI (the Duodenum and upper Jejunum), known as the ‘absorption window’^3,30,32–35^. However, Caramago et al. ^30^ and Flashner et al.^35^ provided empirical evidence that levodopa absorption is comparable along the entire SI. The location of levodopa absorption within the small intestine is important because it affects the duration of absorption and therapeutic effects. We formulated two models to test the two hypotheses. Model A accounts for an ‘absorption window’, Levodopa is effectively absorbed only from the upper third of the SI. In this model, the [effective absorption] term takes the form,

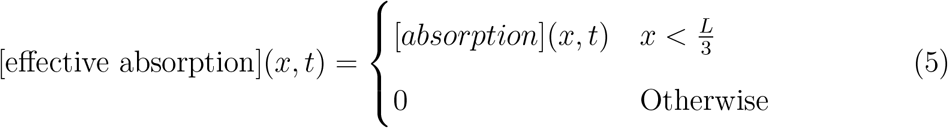

Where [absorption] represents the absorption of levodopa by the LNAA transport system (as described in Equation 14 in the Methods), and *L* is the length of the SI. Alternatively, model B, accounts for an effective absorption along the entire SI. In this model, the [effective absorption] term takes the form,

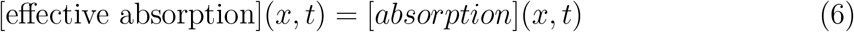

The unabsorbed levodopa and the undegraded CRF in the distal end of the SI, eventually pass into the colon, from which the absorption is negligible^15^.

#### Blood

The concentration of levodopa in the blood (Figure 1d) is determined by the uptake of dissolved levodopa along the SI, and the elimination and the distribution processes in the body^36,37^. Following^37^, we describe the elimination and distribution processes of levodopa in the body using a single-compartment model^38^. The ordinary differential equation describes the kinetics in the concentration of levodopa in the blood,

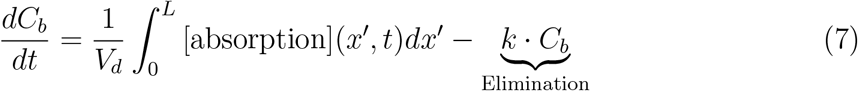

Where *V*_*d*_ represents the volume of distribution of levodopa, *k* is the elimination rate from the body, and *C*_*b*_ is the blood concentration of levodopa.

### Testing model A and model B

Before we use the model, it is necessary to ascertain its ability to predict the overall absorption process of levodopa in healthy and parkinsonian subjects following administration in a commercially available CRF. Due to the conflicting data in the literature on the absorption sites of levodopa along the SI, we developed two models: one that considers an ‘absorption window’ (model A) and one that considers absorption along the entire small intestine (SI) (model B). We now test the two models by comparing their predictions to the blood concentrations of levodopa obtained following oral administration of levodopa in a CRF.

We begin by comparing the model predictions to the experimental blood concentrations of levodopa following administration of Sinemet CR 200mg to healthy fasting subjects (Figure 2a black circle line, taken from Arav^16^). The experimental concentrations reach a maximum within 3 hours and show a double peak. The characteristics of the mean concentrations reflect the kinetics of the individuals as the rise, the location, and the amplitude of the double peaks and the decay are similar to those observed in the individuals (see supplementary information for further details).

**Figure 2.**
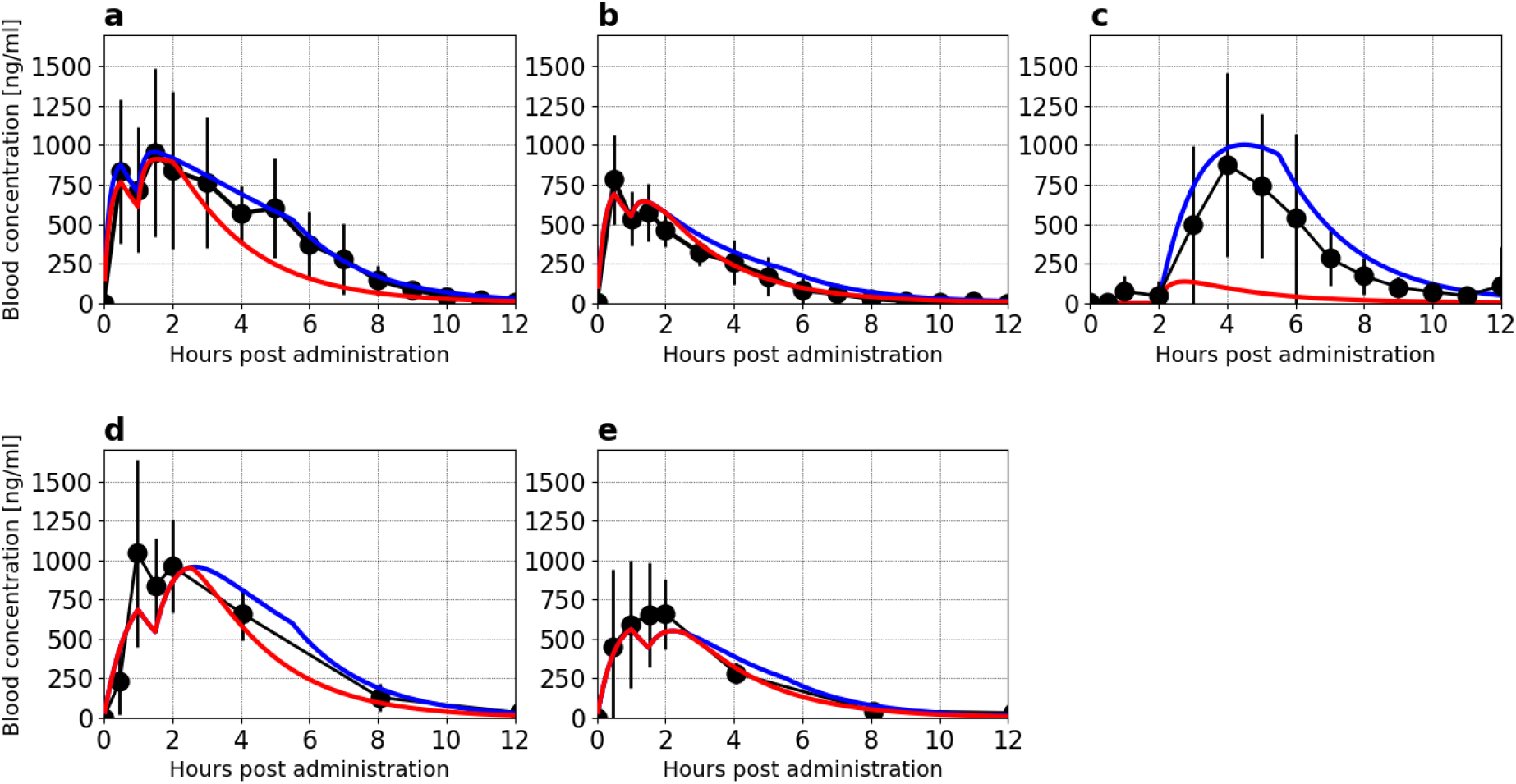
Comparing experimental (black circle line) and the prediction of model A (red solid line) and model B (blue solid line) of blood concentration of levodopa. (a) Sinemet CR 200/50mg in 7 healthy fasting subjects, (b) Sinemet CR 100/25mg in 8 healthy fasting subjects, (c) Enteric-coated controlled release 200mg in 12 healthy fasting subjects, (d) Sinemet CR 200/50mg in 12 parkinsonian fasting subjects and (e) Sinemet CR 100/25mg in 12 parkinsonian fasting subjects. Data for Sinemet CR in healthy and Parkinsonian patients was taken from ^16^, and ^39^, respectively. Data for enteric-coated controlled release was taken from^19^.

To test the model prediction for Sinemet CR 200mg, we solve the equations of model A and of model B using parameters that were compiled from various sources in literature^12,13,23,25,26,29,35,40–43^ (Table S1, supplementary information). Figure 2a compares the predictions of Model A (red) and Model B (blue) to experimental data. As shown, the predictions of both models align well with the experimental mean blood concentrations during the first 2 hours after administration, capturing the rise, location, and magnitude of the two peaks. However, Model A underestimates the concentration of levodopa in the blood starting 2 hours after taking Sinemet CR 200mg, while model B captures the kinetic in the blood well. The overall absorption (bioavailability) of Sinemet CR 200mg is under-predicted by Model A, while the prediction of Model B is within the range reported in the literature. Specifically, the predicted bioavailability of Model A and Model B is 51% and 77%, respectively, and the reported range is 71% to 78%,^18,44^). These results suggest that a substantial portion of Sinemet CR 200mg, approximately 20 *−* 25%, is absorbed in the lower two-thirds of the SI.

Figure 2b depicts the experimental (black circle line, taken from Arav^16^) and the predictions of model A (red solid line) and B (blue solid line) following administration of Sinemet CR 100mg to healthy fasting subjects. The mean concentration profile reflects the kinetic patterns observed in the individual subjects (see supplementary information for further details). As illustrated, both models align well with the experimental data, capturing the temporal patterns, peak locations, and decay of levodopa in the bloodstream. As seen, both models capture well the kinetics of the levodopa in the blood. Specifically, the rise, the location of the peaks, and the decay. This finding suggests that a negligible portion of Sinemet CR 100mg is absorbed in the lower two-thirds of the SI. Figure 2c depicts the experimental (black circle line, taken from Flashner et al.^35^) and the predictions of model A (red solid line) and B (solid blue line) following administration of enteric-coated controlled release (ECCR) reported in Flashner et al.^35^. The ECCR was designed to release levodopa only in the lower Jejunum and the Ileum. As seen, the measured mean blood concentration profile of levodopa begins its rise 2 hours after administration and exhibits a single peak after 4 hours. The mean concentration profile reflects qualitatively the kinetic patterns observed in the individual subjects (see supplementary information for further details). Model A and Model B predictions were obtained using the ECCR release term in Equation 1 corresponding to the ECCR tablet (see Methods for further details). Model A underestimates levodopa’s experimental blood concentrations, while Model B aligns well with the experimental data. These results prove that levodopa is absorbed well along the entire SI.

The results above demonstrate that the proposed model accurately predicts the concentration-time profile of levodopa in the blood following administration of Sinemet CR to healthy subjects. However, previous studies have reported that patients with Parkinson’s disease (PD) experience stomach emptying and intestinal motility delays, which may affect levodopa absorption’s kinetics^36,45–47^. Other pharmacokinetic aspects of levodopa are not impacted by PD^36,45–48^. To investigate this, we compared the experimental results to the model predictions of the concentration-time profile of levodopa following the administration of Sinemet CR to parkinsonian subjects. The mean blood concentrations of levodopa following administration of Sinemet CR 200mg to parkinsonian patients, as reported in Hammerstad et al.^39^, are depicted in Figure 2d (black circle line). As seen, the mean blood concentration of levodopa in parkinsonian subjects is similar to that in healthy subjects (compare to Figure 2b) with similar rise, location, and amplitude of the two peaks and decay. To predict the mean blood concentration of levodopa in parkinsonian subjects, we modified the model by lengthening the residence time of the CRF in the stomach and slowing its emptying rate (see Table S1 for details). The predictions of Model A and Model B are depicted in Figure 2d (red and blue solid lines, respectively). Both models capture the main features of levodopa’s kinetics in the blood of parkinsonian patients, including the rise and decay kinetics and the location of the double peak. Both models slightly underestimate the mean concentration of levodopa in the blood 1 hour after taking Sinemet CR 200mg. However, the measured blood concentrations exhibited a large standard deviation during that time, indicating that a highly variable process determined the blood concentrations. Model A predicts a slightly faster decline in the concentration of levodopa in the blood 2 hours after taking Sinemet CR 200mg than model B. Nevertheless, both models provide a good description of the data. These results also suggest that a smaller fraction of the dose is absorbed in the lower SI of parkinsonian subjects compared to healthy subjects.

Figure 2e depicts the experimental results (black circle line, taken from Hammerstad et al.^39^) following the administration of Sinemet CR 100mg to parkinsonian subjects. The maximal blood concentrations are obtained within 2 after administration. The concentrations are comparable to the concentrations obtained following administration to healthy subjects (Figure 2b, black circle line), albeit the results does not exhibit a double peak. However, exhibit significant inter-individual variation. Nevertheless, the predictions of Model A (Figure 2e, red solid line) and Model B (Figure 2e, blue solid line) capture well the kinetics of the measured levodopa blood concentrations.

We conclude that levodopa is absorbed well along the entire SI and does not exhibit an ‘absorption window’ and that Model B predicts the kinetics of levodopa in the blood following a CRF for healthy and Parkinsonian patients. Hence, we will refer only to Model B when developing the guidelines in the following.

### Guidelines for the design of an optimal CR formulation

As a first step in developing the guidelines, we use the model to understand how the properties of Sinemet CR 200mg influence levodopa blood concentrations in Parkinsonian patients (Simulation 2d) and how they impact the control of PD symptoms. This analysis will allow us to identify the necessary improvements to optimize the CRF.

We first recall that the concentration of levodopa in the blood is determined by the balance between the overall absorption from the SI ([effective absorption] term, Equation 6) and the elimination term ([elimination] term, Equation 7). The [effective absorption] term is determined by the concentration of dissolved levodopa, which in turn depends on the release rate of levodopa from Sinemet CR and the emptying rate of dissolved levodopa from the Stomach.

Using the model, we calculated the release rate of levodopa from Sinemet CR, the emptying rate of dissolved levodopa from the stomach, and the absorption rate from the entire SI into the blood (black, green, and red square lines, respectively in Figure 3). During the first 1.5 hours after administration, the absorption of levodopa is determined by the stomach’s emptying rate (compare the green and red square lines, Figure 3d). Therefore, during that time, the erratic gastric emptying leads to pulsatile blood concentrations. Between 1.5 to 3 hours after administration, the absorption rate of the levodopa is influenced by both the rate at which the stomach empties and the release rate of Sinemet CR. After 3 hours, Sinemet CR reaches the Ileum, and at that time, the absorption of the drug is mainly controlled by the release rate of Sinemet CR (compared to the black line and red square lines).

**Figure 3.**
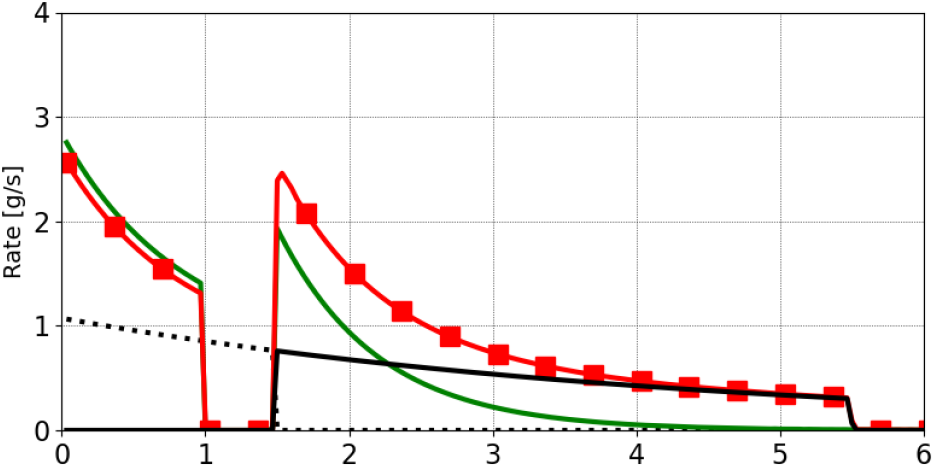
Predicted absorption rate of levodopa following administration of Sinemet CR 200mg to parkinsonian patient (Simulation 2d). The rate of Sinemet CR release rate in the Stomach (black dashed line) and the SI (black solid line), the [release] term in Equation 1; transfer from the stomach to the SI (green solid line), the [Stomach emptying] term in Equation 2; and uptake from the SI to the blood (red square line), the [effective absorption] term in Equation 6.

Two points can be drawn from the results of Figure 3:

i. Rapid release of levodopa in the stomach produces a fast significant increase in levodopa blood concentrations but could result in pulsatile blood concentrations. These are considered to be one of the key factors that trigger the development of motor complications ^1,6,7^.
ii. The release rate of levodopa from the CRF primarily controls the absorption rate of levodopa that was released in the SI. Hence, maintaining a constant release rate throughout the entire SI would lead to constant blood concentrations of levodopa. The release rate should be adjusted to stabilize the blood concentration of levodopa within the therapeutic range.

Based on the two points discussed earlier, it is clear that releasing levodopa in the stomach affects the blood concentration kinetics differently than releasing it in the SI. Therefore, we characterize the putative CRF by the amount of levodopa it releases in the stomach and the amount and release rate in the SI. We use *D*_*St*_ to denote the dose of levodopa released in the stomach and *D*_*SI*_ to denote the dose released in the small intestine.

The characteristics of the CRF (*D*_*St*_, *D*_*SI*_, and release rate) determine the kinetics of levodopa in the blood. Therefore, it is first necessary to identify the target blood concentrations. As previously discussed, control of PD symptoms can be achieved when blood concentrations of levodopa are within the therapeutic range; thus, maintaining a blood concentration sufficient to alleviate PD symptoms while not high enough to cause motor complications, such as dyskinesia^49^. However, the therapeutic threshold for levodopa blood concentrations varies according to the progression stage of PD, and it is commonly related to the Hoehn and Yahr (H&Y) clinical stage^15^. The target blood concentration was set to be the mean of the therapeutic threshold and the dyskinesia threshold for each H&Y stage, and they are summarized in Table I.

**Table I.** The efficacy threshold values for the different disease progression stages on Hoehn and Yahr clinical scale (HY1 to HY4)^15^ and the properties of the CRF that maintains concentrations.

| Stage | Therapeutic window ($ng \cdot ml^{-1}$) | Target Concentrations ($ng \cdot ml^{-1}$) | Release rate ($mg \cdot min^{-1}$) | Dose ($mg$) |
| --- | --- | --- | --- | --- |
| I | 200 – 800 | 400 | 0.19 | 70 |
| II | 300 – 800 | 500 | 0.25 | 90 |
| III | 600 – 800 | 700 | 0.35 | 125 |
| IV | 790 – 800 | 800 | 0.38 | 140 |

The properties of the optimal CRF are generally determined by the initial levodopa blood concentrations of the patient before administration. Specifically, whether the blood levels of levodopa are far below the therapeutic threshold or only slightly below it. In the case of low initial levodopa blood concentrations, typically representing the first dose of the day, it is necessary to increase it significantly and rapidly. This can be achieved by releasing a portion of the dose in the stomach, followed by a slow release in the SI to maintain therapeutic concentrations. In the case of moderate initial levodopa blood concentrations, typically representing the doses throughout the day, it is sufficient to release levodopa at a slower rate to adjust and maintain levodopa blood concentrations within the therapeutic range.

The guidelines for the case of moderate initial blood concentrations will be presented below, as it is more common. The guidelines for an optimal CRF when the initial blood concentrations are low are detailed in the supplementary information.

First, since the patient’s levodopa blood concentrations are moderate, there is no need to quickly elevate levodopa blood concentrations after administering the medication. Hence, it is preferable to avoid releasing levodopa in the stomach (that is, *D*_*St*_ is 0*mg*) since it causes erratic gastric emptying and leads to pulsatile blood concentrations. However, delayed absorption for one or two hours can cause issues, as some patients do not believe the dose has worked and consequently over-medicated themselves^4^. One possible solution is using a multi-particulate extended-release formulation, such as Rytary^50^. In these formulations each particle can be coated with an enteric coating that would prevent them from releasing levodopa in the stomach, but they are not retained in the stomach due to their size^51^.

Since the CRF releases levodopa only in the SI, the rate of levodopa release and elimination govern levodopa blood concentrations (Figure 7, and replacing the absorption term with the release rate). Therefore, the release rate that would maintain the blood concentrations at *C*_*target*_ is given by (see supplementary information for further details),

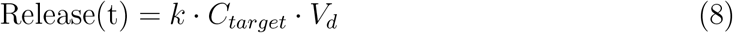

The target blood concentrations and the release rate required for each stage are presented in Table I. The dose (*D*_*SI*_) is calculated based on a total transit time of 6 hours. The stomach emptying rate in parkinsonian patients is approximately 1.5 hours, the intestinal transit time is at least 3 hours^29^, and an additional residence time of 1.5 hours in the terminal ileum before emptying to the colon^42^. It is expected that, on average, the putative CRF would maintain stable therapeutic concentrations for around 6 hours.

## Discussion

Developing a CRF that would deliver a consistent and sustained therapeutic blood concentration of levodopa proved to be a difficult task^3^. To facilitate the development of such formulation, we developed a physiologically-based mathematical model that describes the absorption of levodopa following administration in a CRF (Equations 1 to 7). Our key results: (i) levodopa is effectively absorbed throughout the entire SI (ii) the release of levodopa in the stomach causes fluctuations in the blood concentrations during the first 3 hours after administration; (iii) inadequate levodopa release in the ileum shortens the duration of the therapeutic effect of controlled-release formulations (iv) the ideal release rate, and dosage are contingent on the stage of PD.

Previous studies ^3,32–34^ have suggested that levodopa absorption is primarily restricted to the upper third of the SI (i.e exhibits an ‘absorption window’). However, Caramago et al.^30^ have shown that the levodopa transporter is expressed along the entire SI of mice. Flashner et al.^35^ examined levodopa uptake in the distal SI by administrating a controlled release enteric coated formulation designed to begin its release in the Ileum. Their results indicate that levodopa is well absorbed in the Ileum. The model’s ability to accurately predict the mean blood concentrations of healthy and parkinsonian individuals (Figure 3) further supports the hypothesis that levodopa is absorbed well along the entire SI.

The physiologically-based mathematical model developed in this study describes the kinetics behavior of levodopa absorption following its administration in a CRF (Figure 1). The parameters that describe the physiological processes were derived from in-vivo studies^12,13,23,25,26,29,40–42^, while the release rate from the CR formulation was taken from in-vitro experiments^35,43^. This opens the possibility of using the model during the research and development phase of the formulation, thereby reducing the number of clinical trials.

We have shown that shifting part of the release of levodopa from the Stomach to the Ileum is expected to reduce fluctuations in blood concentrations of levodopa, increase the bioavailability and prolong the therapeutic effect to 6-8 hours. This estimation is based on the SI transit time of a CRF in healthy subjects. Patients with PD, especially in later stages, often suffer from constipation, which might cause prolonged transit time of formulation in the SI^52,53^. Longer transit time may result in prolonged maintenance of levodopa in the therapeutic concentrations of levodopa^48^.

Developing a CRF that would implement the guidelines detailed in this study would require additional pharmaceutical effort. A crucial step is the selection of the pharmaceutical dosage form. Using a single-unit enteric coated formulation, similar to the one described in a study by Flashner, et al.^35^, is straightforward as this technology is well established. However, monolithic tables remain in the stomach for 1 *−* 1.5 hours^29^, and possibly even longer for patients with PD, so the relief from PD symptoms will be delayed. This might pose a problem since some patients might not believe that the dose worked and take another dose^4^. A possible solution to this problem is to use enteric-coated multi-particulate delivery systems ^51,54^, such as the extended-release formulation of levodopa IPX066 (Rytary)^55^. Such formulations comprised of beads (or pellets) with diameters under 2mm with a coating that releases levodopa at different rates along the SI. Particles of such size can pass the pylorus continuously^51^, and therefore elevate the blood concentrations of levodopa shortly after the administration.

Another key finding is that the tablet erosion rate determines levodopa uptake into the blood once the CR formulation reaches the SI, and no dissolved levodopa remains in the stomach. We used this information to calculate the desired erosion rate for the different stages of PD that would lead to therapeutic levels, as shown in Table I. Targeting the different stages of PD is beneficial because it allows for lower therapeutic concentrations to be reached in the early stages (H&Y1 and H&Y2), reducing the need for large doses of levodopa. This finding also implies that the dissolved levodopa is exposed to degrading processes (the microbiota, luminal enzymes, and chemical reactions^15^) in the SI for a short period. Therefore, these processes’ effect on the overall Sinemet CR bioavailability is expected to be minimal.

One limitation of this study is that it does not consider a meal’s influence on absorption kinetics. The presence of a meal can delay stomach emptying and increase competition between the amino acids in the meal and levodopa for transport across the gut, brain, and kidneys, as described in a study by Guebila et al.^15^. The impact of meals on levodopa absorption has been studied in both experimental^26,48,49,56^, and theoretical^15^ studies.

In conclusion, we demonstrated that adjusting the release rate of a CR formulation can potentially extend the therapeutic duration of a single dose of levodopa. This supports using a combination of theoretical modeling, in-vivo experiments, and in-vitro testing as a powerful tool for developing novel CR formulations.

## Methods

### Mathematical model

The physiologically based mathematical model represents the kinetics of the physiological processes that take place during the absorption of levodopa and were delineated in Figure 1. In this section, we provide a detailed description of the mathematical model, which is represented by Equations (Equations 1 to 7).

The release rate of levodopa from the Sinemet CR formulation is described by Equation 1. To complete this equation, it is necessary to specify the details of the Erosion term. The erosion rate of Sinemet CR in vitro follows a first-order profile^43^. That is, the erosion rate is proportional to the amount of drug remaining within the system and thus declines with time. Hence, the erosion term is,

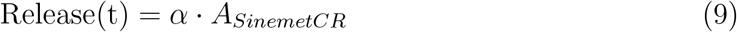

Where *α* (*s*^*−*1^) is the erosion rate coefficient. The rate coefficient was calculated by fitting the solution to equation 1 to the erosion rate that was reported in vitro^43^. It has been reported that in many monolithic matrix controlled release tablets, such as Sinemet CR, there is an initial large release of the drug (‘burst release’) before the release rate reaches a stable profile. This phenomenon is often observed in the literature^43,57^, and hence we have incorporated such a process into the model.

The transfer rate of dissolved levodopa from the stomach to the SI is described by equation 2. To complete this equation, it is necessary to specify the stomach emptying term. As described by Oberle et al.^41^, stomach emptying is modeled as a first-order process (that is, the transfer rate is proportional to the concentration in the stomach). When dissolved levodopa is present in the stomach, the stomach emptying process is temporarily halted, resulting in a lag phase during which no transfer occurs^25,26^. Hence we have incorporated such a lag in our model. Specifically,

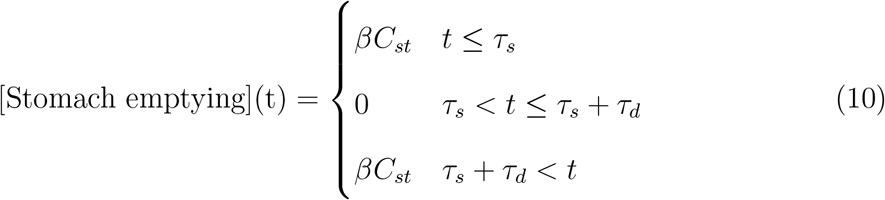

Where *τ*_*s*_ and *τ*_*d*_ are the beginning of the lag in stomach emptying and its duration, respectively.

The processes in the SI are described with equation 3. To complete the description of this equation, it is necessary to describe the transit term that describes the propulsion and mixing of dissolved levodopa along the SI and the term that describes its uptake to the blood. The transit term describes the concomitant propulsion of dissolved levodopa by the SI walls toward its end and the mixing. As was mentioned above, the SI is described as a continuous tube similar to^12,14,24^, but contrary to the compartmental approach^15,58^. The propulsion of the dissolved levodopa toward the distal SI is described by a convection term of the mean velocity, while the mixing along the SI is described as a dispersion term (that is, an effective diffusion term). Specifically,

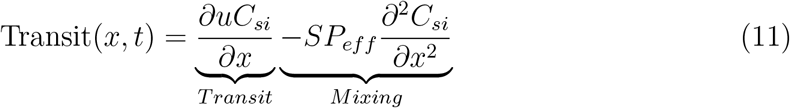

where *u m · s*^*−*1^ is the mean convection velocity and *SP*_*eff*_ *m · s*^*−*2^ is the effective longitudinal mixing rate of the SI. The negative sign of the mixing term denotes that the mixing flux is in the counter-gradient direction. Note that the intestinal equation comprises a 1D partial differential equation, which requires boundary conditions. These boundary conditions describe the flux of dissolved levodopa to the beginning of the SI from the stomach and the flux of dissolved levodopa from the SI to the large intestine at its distal end. Accordingly, The emptying from the stomach and the emptying to the large intestine is described as

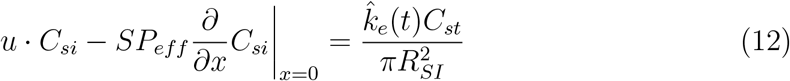

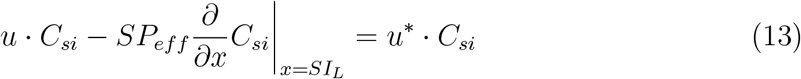

Where *R*_*lumen*_ is the radius of the SI. The contents of the SI reside in the terminal SI for approximately 1.5 hours before it is emptied to the colon^42^. Hence *u*^*∗*^ equals 0 during that time and *u* when the SI is emptied.

The uptake of dissolved levodopa from the SI to the blood is mediated by the L-neutral amino acid transport system for large amino acids (LNAA). Hence, we described the uptake as a saturable process,

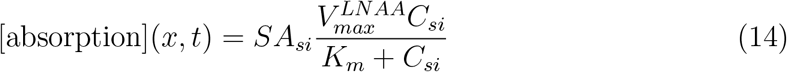

Where *SA*_*si*_ is the amplification of the small intestine surface area due to the folds and the villi^13^, 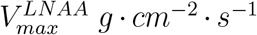 and *K*_*m*_ *g · cm*^*−*3^ are the maximal transport rate and the association constant of levodopa across the epithelial boundary layer.The kinetics of levodopa blood concentrations are described in equation 7. To complete the description of this equation, the elimination term must be described. The elimination of levodopa from the body follows first-order kinetics^37^. Therefore,

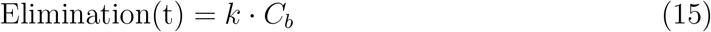

Where k (*s*^*−*1^) is the elimination rate coefficient, and *C*_*b*_ (*g ·cm*^*−*3^) is the levodopa concentration in the blood. The elimination term reflects the fact that levodopa is eliminated from the body by various mechanisms such as metabolism, excretion, and degradation.

### Numerical methods

The model’s equations were solved numerically using in-house software (available at https://github.com/yehudarav/levodopaAbsorption). The software was developed using the open Field Operation and Manipulation (OpenFOAM) framework (version 7)^59^, which extends the C++ programming language to include tensor calculus. The temporal derivations were solved using a first-order implicit scheme. The partial differential equation (Equation 3) was solved using the finite volume method.

## Data Availability

All data produced in the present study are available upon reasonable request to the authors

## Author Contributions

YA - conceptualization, programming, analysis, writing. AZ - analysis,writing.

## Acknowledgement

The authors wish to thank Prof. Hanna Parnas for her insights and comments.

## Supplemental Information

Supplementary Information is available for this paper.

