## Supplementary information for "Model-based optimization of controlled release formulation of levodopa for Parkinson’s disease"

#### Parameter selection and model validation

To validate the model, we compare its predictions to experimental observations. This validation process was performed in two stages: first, we compared the model predictions to the observed blood concentration of healthy subjects after administering Sinemet CR and an enteric-coated controlled release formulation. This step is important as the rate constants used in the model were derived from studies involving healthy individuals. In the second stage, we demonstrate that the model accurately predicts the observed blood concentrations in individuals with Parkinson's disease by using slower rate constants for gastric emptying and intestinal motility.

##### Healthy subjects

Sinemet CR 200/50mg (200 mg levodopa and 50mg Carbidopa, an AADC) was administrated to 7 non-smoking healthy male subjects (ages 18-55) [1]. The subjects fasted for at least 10 hours before administration. Figure 2a depicts the mean blood concentration-time profile of levodopa (The plasma kinetics of the individuals is depicted in Figure S2). The maximal concentrations and a double peak are obtained during the first 3 hours after administration. At later times, the concentration of levodopa in the blood decays with a half-life time of approximately 1.5 hours, which is the characteristic decay rate of levodopa following concomitant administration with AADC inhibitor [2]. The occurrence of a double peak in blood concentrations following oral administration of levodopa is well documented in the literature[3–5] and observed in 5 out of 7 subjects (subjects 1,2,3,6 and 7, Figure S2) following administration of Sinemet CR 200/50mg.

We also examined the model predictions following the administration of Sinemet CR 100/25mg (100 mg levodopa and 25mg Carbidopa) to 8 healthy fasting volunteers (Figure 2b, the blood concentration-time profile of the individuals is depicted in Figure S1). The mean blood concentrations of levodopa, depicted in exhibit similar behavior to Sinemet

CR 200/50mg; the maximal concentrations obtained in the first 2 hours, a double peak with a second small peak, and the half-life time of the decay is 1.5 hours. Double peak is observed in 4 out of 8 following administration of Sinemet CR 100/25mg (subjects 1,3,4 and 7, Figure S1).

The model predictions were obtained by solving equations 1 to 7. All the model parameters were compiled from various sources in literature[4–14] (Table S1), except for the dose dumped ('burst'). This parameter was obtained by fitting to the observed blood concentrations of healthy subjects at 0.5h. The value of $V_{max}^{LNAA}$ was obtained by fitting the model results to the data of Gundert-Remy et al.[15] that infused levodopa solution to the Duodenum.

**Table S1.** The parameters used in the levodopa simulations.

| Parameter Name | Value | Reference | Comment |
|---|---|---|---|
| **Stomach** | | | |
| Volume ($V_{st}$) | 200ml | [11] | |
| lag duration ($\tau_d$) | 30min | [5] | |
| *Healthy subjects* | | | |
| $t_{1/2}$ | $5min$ | [11] | |
| lag start ($\tau_s$) | $30min$ | [5] | |
| stomach residence time | $1h$ | [7,16] | |
| *Parkinsonian subjects* | | | |
| $t_{1/2}$ | $30min$ | | See text |
| lag start | $1h$ | | See text |
| stomach residence time | $1.5h$ | | See text |
| **Small Intestine (SI)** | | | |
| Length ($L_{si}$) | 282cm | [17] | |
| Radius ($R_{si}$) | 1.5cm | [6,17] | |
| Transit velocity ($u_{si}$) | $2.6 \cdot 10^{-2}\ cm \cdot sec^{-1}$ | [18] | 3 hours |
| Spread coefficient ($SP_{si}$) | $0.49\ cm^2 \cdot sec^{-1}$ | [18] | |
| Terminal residence time | 1.5 hours | [12] | |
| Surface Area amplification ($SA_{si}$) | | [6] | |
| LNAA $V_{max}$ ($V_{max}^{LNAA}$) | $1.65 \cdot 10^{-6}\ g \cdot s^{-1} \cdot cm^{-2}$ | | See text |
| LNAA $K_m$ ($K_m^{LNAA}$) | $7.5mM$ | [9] | |
| **Body** | | | |
| Volume of distribution ($V_d$) | 63L | [2] | $0.9\ L \cdot kg^{-1}$ |
| Elimination rate constant ($k_{10}$) | $1.28 \cdot 10^{-4}\ s^{-1}$ | [10] | $t_{1/2}$ of 1.5 hours |
| **Formulation** | | | |
| *Sinemet CR* | | | |
| Commence of release | $0h$ | [14] | |
| first order release $t_{1/2}$ | $3h$ | [14] | |
| dose dump ('burst') | $60mg$ | See text | |
| *Enteric coated CR* | | | |
| Commence of release | $2h$ | | See text |
| first order release $t_{1/2}$ | $3h$ | | See text |
| dose dump ('burst') | $0mg$ | | See text |

The model predictions for Sinemet CR 200/50mg and 100/50mg are shown in Figure 2a and b (respectively). As seen, the model captures well the kinetics of the blood concentrations of levodopa. Specifically, the model describes the peak's location, magnitude, and decay.

To further test the model, we also compared it to the experimental results obtained by administrating an Enteric-Coated Controlled-Release (ECCR) levodopa formulation (200mg). This formulation was developed by Teva R&D Initiative[13] and was administrated with 2x25mg Carbidopa to 12 healthy volunteers. The ECCR formulation was designed for beginning its release in the first third of the SI (the end of the Jejunum). The erosion rate of the ECCR formulation was measured in-vitro and found to be approximately first order with $t_{1/2}$ of 2 hours[13] and given in Table S1.

##### Parkinsonian subjects

To predict the mean blood concentrations of levodopa for parkinsonian patients, it is necessary to estimate how the constant rates of the different processes in the model vary concerning a healthy person. Previous studies[19–22] have shown that rate constants for these processes are similar for healthy and parkinsonian patients with the exception that stomach emptying and intestinal motility are retarded by PD. Hence, we have adjusted the model parameters determining gastric emptying and gastrointestinal motility: the stomach emptying $t_{1/2}$ and the residence time of Sinemet CR in the stomach were fitted to the experimental results (Figure 2). The values of the fitted parameters are given in Table S1.

#### Sinemet CR 100mg individuals

In this section, we describe the blood concentrations of healthy individuals following the administration of Sinemet CR 100mg taken from Arav et al.[1]. Figure S1 shows levodopa blood concentration in 8 individuals following administration of Sinemet CR 100/25mg (100mg levodopa and 25mg Carbidopa, an Aromatic L-Amino acid Decarboxylase). The individuals were non-smoking healthy male subjects (ages 18-55)[1], and fasted for at least

10 hours before administration.

In all the individuals, the peak levodopa blood concentrations are obtained within the first hour after administration. In 4 of the 8 individuals (numbers 1,4,5, and 8), the concentration-time profile exhibits a double peak, where the second peak is smaller than the first. In 7 of the 8 individuals (numbers 1,2,3,4,5,7, and 8), the concentrations decay smoothly after 2 hours. Individual 6 exhibited a small rise in the blood concentrations after 4 hours before the concentrations decayed. These results show that the rise, peak magnitude and location, and decay of the mean levodopa blood concentrations are similar to those observed in the individuals. Therefore, we can conclude that the mean concentrations of levodopa in the blood accurately represent the kinetics of levodopa in the individuals.

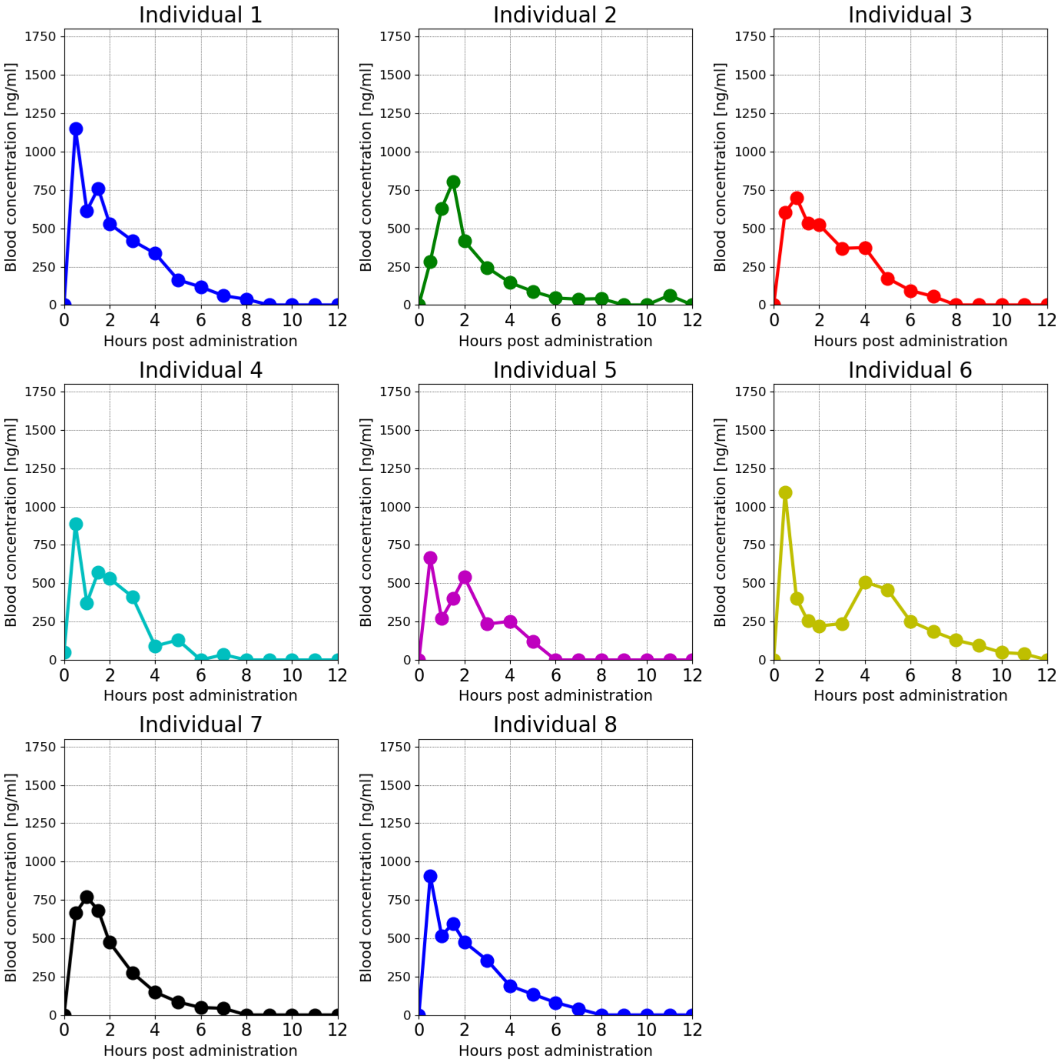


**Figure S1.** Experimental Concentration time profile following administration of Sinemet CR 100/50mg to healthy subjects. Taken from[1].

### Sinemet CR 200mg individuals

In this section, we describe the blood concentrations of healthy individuals following the administration of Sinemet CR 200mg taken from Arav et al.[1]. Figure S2 shows levodopa blood concentrations in 7 individuals following administration of Sinemet CR 200/50mg (200 mg levodopa and 50mg Carbidopa, an Aromatic L-Amino acid Decarboxylase). The individuals were non-smoking healthy male subjects (ages 18-55)[1], and fasted for at least

10 hours before administration. The concentration-time profiles of levodopa in 5 subjects (individuals 1,2,3,6,7) exhibit similar behavior. Specifically, the maximal concentrations and double peaks were seen in the first 3 hours. The other 2 subjects (individuals 4 and 5) exhibit a delayed and slow absorption, with maximal concentrations obtained 4 to 5 hours after administration. In all subjects, the half-life time of the decay of levodopa in the blood is 1.5 hours, which is the characteristic decay rate of levodopa following concomitant administration with AADC inhibitor[2]. The occurrence of double peaks in blood concentrations following oral administration of levodopa is well documented in the literature and was attributed to an erratic gastric emptying[3–5].

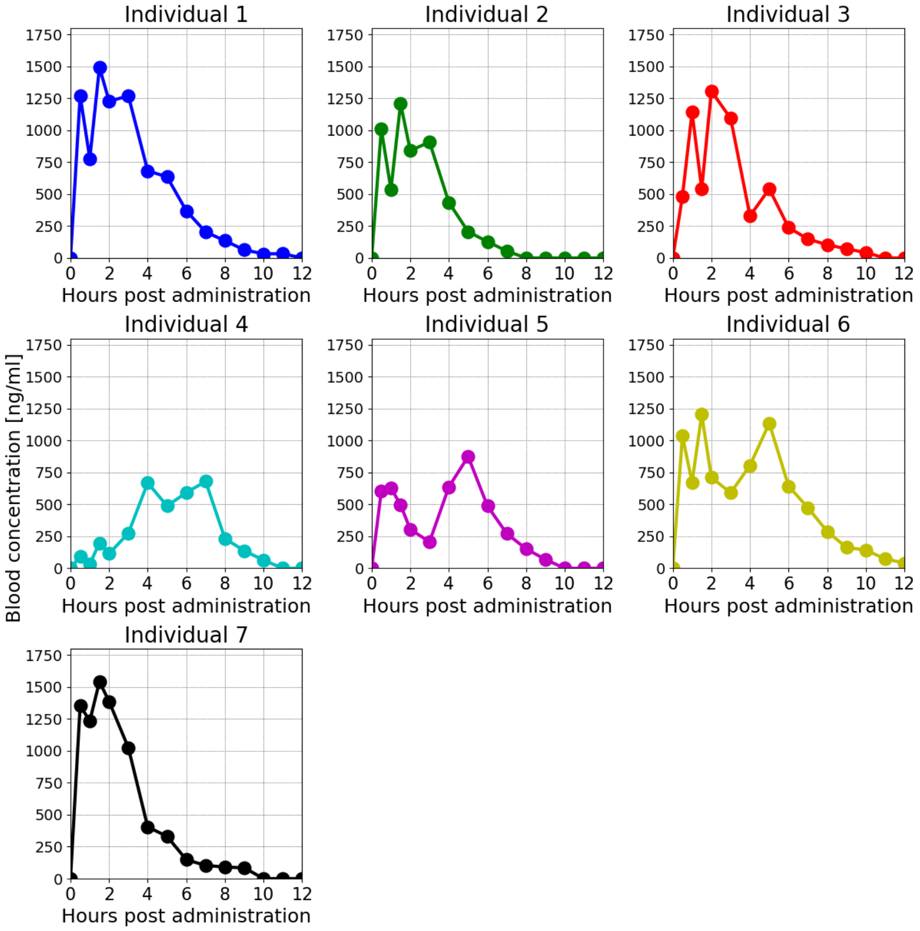


**Figure S2.** Experimental concentration-time profile after administering Sinemet CR 200/50mg to fasting subjects. (a-g) concentration-time profile of healthy individuals, Data of healthy individuals were taken from[1].

These results show that the rise, peak magnitude and location, and decay of the mean levodopa blood concentrations are similar to those observed in the individuals. Therefore, we can conclude that the mean concentrations of levodopa in the blood accurately represent the kinetics of levodopa in the individuals.

### Enteric coated controlled release 200mg individuals

In this section, we describe healthy individuals' blood concentrations after administering 200mg Enteric-coated controlled release formulation taken from Flashner et al.[23].

Figure S3 shows the concentration-time profile of the 12 individuals. Except for individual 6, the concentration-time profiles of the individuals share a close resemblance to each other. Specifically, the rise in the concentrations begins approximately 2 hours after administration, exhibits a single peak after 4 to 6 hours, and then a decay in the concentrations.

These results show that the rise, peak magnitude and location, and decay of the mean levodopa blood concentrations are similar to those observed in the individuals. Therefore, we can conclude that the mean concentrations of levodopa in the blood accurately represent the kinetics of levodopa in the individuals.

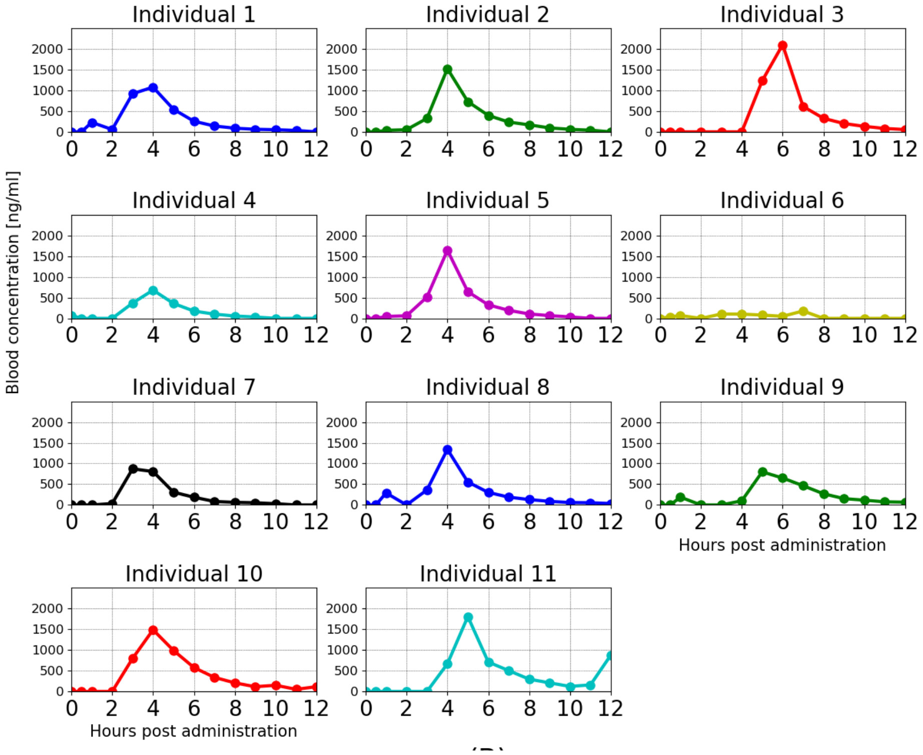


**Figure S3.** Experimental Concentration time profile following administration of Enteric-coated controlled release 200mg to healthy subjects. Taken from[23].

### Finding the optimal dose and the release rate

In this section, we detail the procedure to find the loading and maintenance doses ($D_{St}$ and $D_{SI}$, respectively) and the release rate in the SI. The $D_{St}$, $D_{SI}$, and the release rate are selected to achieve therapeutic blood concentrations and maintain them for as long as possible. Since the therapeutic threshold increases over the course of PD (Table S2), we will select the $D_{St}$ and $D_{SI}$ and the release rate for each stage.

To do so, we recall that the emptying of dissolved levodopa from the stomach and the release from the CRF determine the overall absorption of levodopa into the blood (Figure 3). Hence, the kinetics of levodopa blood concentrations can be calculated by replacing the Absorption term in the blood concentration kinetics (Equation 7) with the sum of the stomach emptying and the release from the CRF (Equation 1),

$$\frac{dC_b}{dt} = \frac{\text{Release(t)+[Stomach Emptying]}}{V_d} - \underbrace{k \cdot C_b}_{\text{Elimination(t)}} \tag{S1}$$

Where the elimination term from the body was taken from Equation 9.

The [Stomach Emptying] rate is calculated solving Equations 2 and 10,

$$\text{[Stomach emptying](t)} = \begin{cases} \beta D_{St} \cdot e^{-\beta \cdot t} & t \leq \tau_s \\ 0 & \tau_s < t \leq \tau_s + \tau_d \\ \beta D_{St} \cdot e^{-\beta \cdot \tau_s} \cdot e^{-\beta \cdot (t-(\tau_s+\tau_d))} & \tau_s + \tau_d < t \end{cases} \tag{S2}$$

Where $\beta$ is the stomach emptying rate, and $\tau_s$ and $\tau_d$ are the beginning of the lag in stomach emptying and its duration in parkinsonian patients, respectively.

The $D_{St}$, $D_{SI}$, and the release rate are determined as follows. First, we note that after the emptying of dissolved levodopa from the stomach ($t \gg (\tau_s + \tau_d)$), the release rate from the CRF is the only source of levodopa in the blood (Term [Release(t)] in Equation S1). Therefore, the release rate that is required to maintain the target concentration $C_{target}$ is

$$\text{Release(t)} = k \cdot C_{target} \cdot V_d \tag{S3}$$

Where $C_{target}$ in each stage was taken to be the mean of the therapeutic and dyskinesia thresholds as the target blood concentration (Table S2).

The value of the $D_{SI}$ is calculated from the release rate and the formulation's transit time in the SI. Since the optimal release rate is constant, the $D_{SI}$ is found by

$$D_{SI} = Release \cdot [Transittime] \tag{S4}$$

The transit time in the SI is at least 3 hours[7], and an additional residence time of 1.5 hours in the terminal ileum before emptying to the colon[12]. However, a multi-particulate formulation, such as Rytary[24], is released slowly from the stomach, and therefore the transit time of multi-particulate formulations is longer. We estimated the time scale of stomach emptying to be approximately 1.5 hours in parkinsonian patients based on the results in Figure 2d. Hence, the total transit time in the SI is approximately 6 hours.

A multi-particulate formulation that releases levodopa at the [Release] rate would maintain levodopa blood concentrations at the desired level. However, sometimes it is necessary to elevate the blood concentration from very low to therapeutic levels. The portion of the dose that was released in the stomach is emptied to the SI and absorbed quickly. Hence, we select the $D_{St}$ to relieve PD symptoms approximately 30 minutes after administration. To do so, we estimate $\tau_s$ and $\tau_d$ in parkinsonian patients to be 1 hour and 30 minutes, respectively, based on the results in Figure 2d (Table S1). Therefore, we can compute the concentration of levodopa in the blood during the first 30 minutes by substituting the stomach emptying rate in Equation S2, in Equation S1,

$$\frac{dC_b}{dt} = \frac{1}{V_d}\left(\text{Release} + \beta D_{St} \cdot e^{-\beta \cdot t}\right) - \underbrace{k \cdot C_b}_{\text{Elimination(t)}} \tag{S5}$$

Where [Release] is constant. This equation can be solved to obtain an analytical solution of the concentration in the blood as a function of $D_{St}$ and $t$,

$$C(t, D_{St}) = \frac{bD_{St}}{V(b-k)}\left[e^{-\beta t} - e^{-kt}\right] + \frac{R}{Vk}\left[1 - e^{-kt}\right] \tag{S6}$$

The singularity in Equation S6 is a mathematical artifact since in reality, the elimination rate $k$ will not be exactly equal to the emptying rate $\beta$. In order to avoid the singularity, one can either obtain a different analytical solution to Equation S5 when $k = \beta$, or slightly perturb one of the rates.

The $D_{St}$ is selected by finding the $D_{St}$ that minimizes the expression,

$$\min_{D_{St}} \left\{ (C(30min, D_{St}) - C_{target})^2 \right\} \tag{S7}$$

The values of $D_{St}$, $D_{SI}$, and the release rate in each H&Y stage are given in Table S2. The acceptable error column presents the margins of error in the release rate that would maintain the concentration within the therapeutic range of the PD stage therapeutic range. In the case of a multi-particulate formulation, the release rate S1 represents the total release rate from all the particles in the SI.

**Table S2.** The efficacy threshold values for the different disease progression stages on Hoehn and Yahr clinical scale (HY1 to HY4)[25] and the properties of formulation that raises concentration to the therapeutic level and maintains it.

| Stage | Therapeutic window ($ng \cdot ml^{-1}$) | Release rate ($mg \cdot min^{-1}$) | Loading dose ($D_{St}$) ($mg$) | Maintenance dose ($D_{SI}$) ($mg$) |
|---|---|---|---|---|
| I | 200 – 800 | 0.19 | 16 | 70 |
| II | 300 – 800 | 0.25 | 28 | 90 |
| III | 600 – 800 | 0.30 | 68 | 104 |
| IV | 790 – 800 | 0.30 | 68 | 104 |

The predicted blood concentrations for the different H&Y stages are presented in Figure S4. As seen, in the formulation that was designed for stages I and II (Figure S4a and b, respectively), the blood concentrations reach the therapeutic range approximately 30 minutes after administration and maintain it for $6 - 7$ hours using $86mg$ and $118mg$ levodopa. The formulation that was designed for stages III and IV (Figure S4c and d, respectively) results in higher than the threshold and the dyskinesia threshold. This is because the therapeutic levels are high and the release from the stomach is slow. Therefore, $D_{St}$ is relatively large and it increases the concentrations at later times. Nevertheless, the formulation maintains above therapeutic concentrations for 5.5 hours with 172 mg of levodopa.

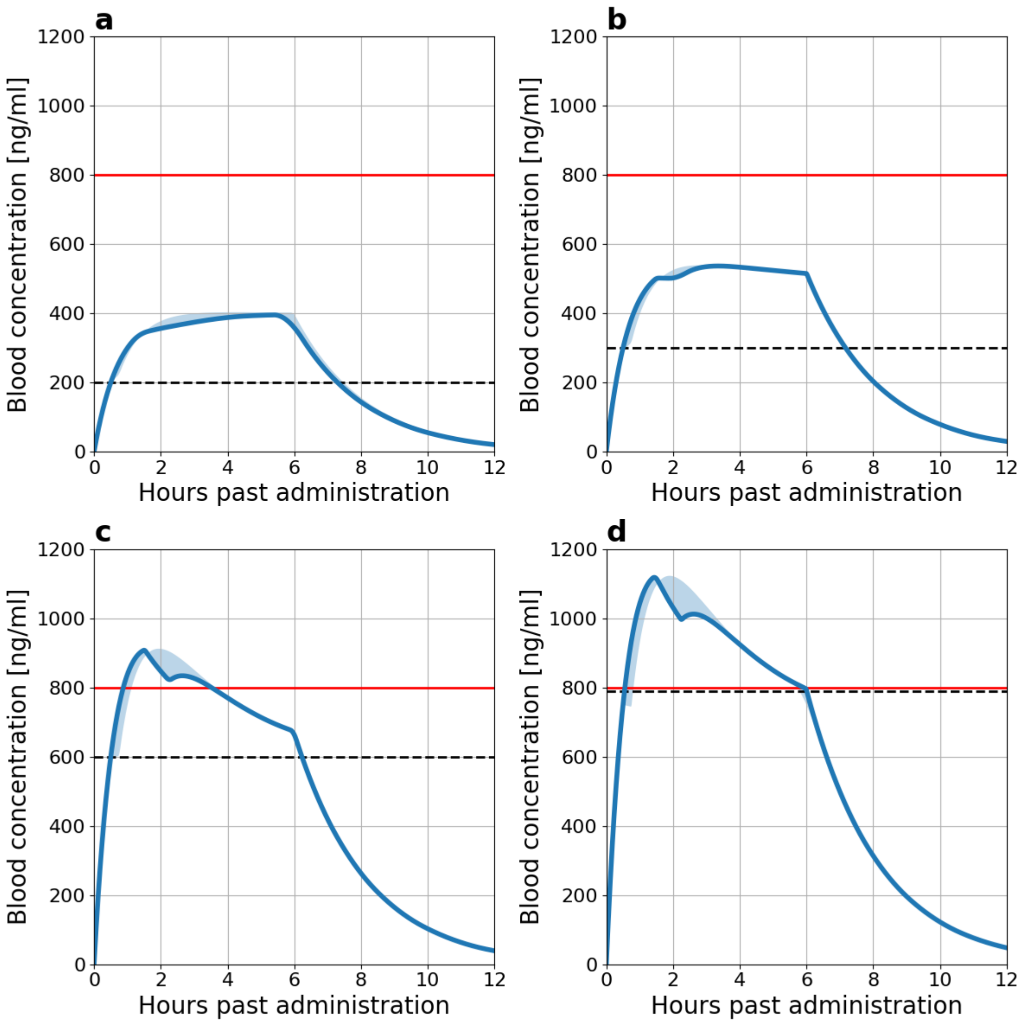


**Figure S4.** The predicted blood concentration of levodopa from the putative CRFs detailed in Table S2. (a) H&Y stage I, (b) H&Y stage II, (c) H&Y stage III, and (d) H&Y stage IV. The shaded areas depict the range of concentration as a result of ±30min in the onset of the lag and ±15min in its duration.
